## Supplementary material for "DORA-compliant measures to assess research quality and impact in biomedical institutions: review of published research, international best practice and Delphi survey": CREDES reporting checklist

CREDES Checklist: Recommendations for the Conducting and REporting of DElphi Studies

| Items | Location in manuscript |
| --- | --- |
| Purpose and rationale. The purpose of the study should be clearly defined and  demonstrate the appropriateness of the use of the Delphi technique as a method to  achieve the research aim. A rationale for the choice of the Delphi technique as the  most suitable method needs to be provided. | Introduction page 6; Methods, Research Design page 6-7 |
| Expert panel. Criteria for the selection of experts and transparent information on  recruitment of the expert panel, sociodemographic details including information on  expertise regarding the topic in question, (non)response and response rates over the ongoing iterations should be reported. | Sampling and recruitment, page 10 |
| Description of the methods. The methods employed need to be comprehensible; this includes information on preparatory steps (How was available evidence on the topic in question synthesised?), piloting of material and survey instruments, design of the survey instrument(s), the number and design of survey rounds, methods of data analysis, processing and synthesis of experts’ responses to inform the subsequent survey round and methodological decisions taken by the research team throughout the process. | Page 6-11 |
| Procedure. Flow chart to illustrate the stages of the Delphi process, including a  preparatory phase, the actual ‘Delphi rounds’, interim steps of data processing and  analysis, and concluding steps. | Figure 2 |
| Definition and attainment of consensus. It needs to be comprehensible to the reader how consensus was achieved throughout the process, including strategies to deal with non-consensus. | Data collection and analysis, page 11 |
| Results. Reporting of results for each round separately is highly advisable in order to  make the evolving of consensus over the rounds transparent. This includes figures  showing the average group response, changes between rounds, as well as any  modifications of the survey instrument such as deletion, addition or modification of  survey items based on previous rounds. | Page 12-17; Tables 1,2 and 3; Appendices S5 to S8, Figure 2 |
| Discussion of limitations. Reporting should include a critical reflection of potential  limitations and their impact of the resulting guidance. | Page 22-23 |
| Adequacy of conclusions. The conclusions should adequately reflect the outcomes of the Delphi study with a view to the scope and applicability of the resulting practice guidance. | Page 23 |

Jünger S, Payne SA, Brine J, Radbruch L, Brearley SG. Guidance on Conducting and REporting DElphi Studies (CREDES) in palliative care: Recommendations based on a methodological systematic review. Palliat Med. 2017;31: 684–706.
