## Supplemental Appendix 1 for "DORA-compliant measures to assess research quality and impact in biomedical institutions: review of published research, international best practice and Delphi survey"

**APPENDIX 1. Scoping review inclusion and exclusion criteria**

**ELIGIBLE**

**Population/People** [studies/documents eligible for review may include one or more of the following participants or targets]

- Independent clinician and non-clinician researchers referred to as investigator, scientist, professor, etc. of any health sciences or life sciences discipline, or candidates for these positions
- Leaders responsible for making hiring, tenure, promotion or other decisions based on researcher productivity or performance (e.g. quantity, quality/excellence, attributes, impact) such as research institutes, directors or vice-presidents of a research portfolio, deans, departmental chairs, etc.
- Based in academic settings including universities, or university-affiliated independent or hospital-based research institutes
- Disciplines chosen from *Canadian Research and Development Classification 2019* (jointly developed by SSHRC, NSERC, CIHR) to represent range of research or research expertise at UHN; for example:

| Group | Class (include many sub-fields) |
| --- | --- |
| Basic medicine and life sciences | Immunology  Cancer  Medical microbiology  Neurosciences  Pharmacology/pharmaceutical sciences  Medical physiology  Musculoskeletal health and human movement |
| Clinical medicine | Cardiorespiratory medicine and hematology  Circulatory diseases  Clinical sciences (e.g. hepatology, surgery)  Pediatrics and reproductive medicine |
| Health sciences | Nutrition  Public and population health (e.g. epidemiology, psychosocial/determinants of health)  Rehabilitation sciences  Health services and systems (catchall for a lot)  Care (family, primary, residential, hospital, nursing, allied/complementary) |
| Medical biotechnology | Gene and molecular therapy  Regenerative medicine  Drug discovery |
| Mathematics/Statistics | Statistics (biostatistical methods) |
| Computer and information sciences | Artificial intelligence  Information systems  Bioinformatics |
| Industrial, systems and process engineering / Materials engineering | Human factors engineering  Systems engineering  Operational research  Materials engineering |
| Medical and biomedical engineering | Biomaterials  Biomedical instrumentation  Medical imaging  Medical devices  Artificial tissues |
| Psychology and cognitive sciences | Psychology  Cognitive sciences  Psychometrics  Judgement and decision-making |
| Sociology | Sociology of healthcare  Sociology and social studies of health, health systems and health care  Social theory  Sociological methodology and research methods  Sociology and social studies of science and technology |
| Education | Higher education  Curriculum and pedagogy theory and development  Medicine, nursing and health curriculum, pedagogy and didactics  Education assessment and evaluation  Inter-disciplinary, trans-disciplinary learning and education |
| History | History of medicine and health care |
| Philosophy | Bioethics  Epistemology and philosophy of sciences |

**Intervention/Issue** [studies/documents eligible for review may focus or mention one or more of the following]

- Assessment of research productivity of individuals, which articles/documents may refer to as impact, performance, etc.
- Productivity may be based on quantitative or qualitative outputs, products, impacts, achievements, ***including but not limited to***:
  - Scientific

National/international recognition, invitations, awards, vetted academy membership (e.g. Canadian Academy of Health Sciences), journal articles, peer-reviewed grant funding, refereed conference proceedings, preprints, books, book chapters, white papers, software, datasets, web servers, invention disclosures, patent applications, issued patents, intellectual property, licenses, commercialization or products, launch of companies, text/video/audio interviews, presentations or podcasts, other knowledge translation modalities, conference abstracts, presentations, posters, etc.

- Health system

Written policies, guidelines or standards, policy/guideline/standard implementation, policy consultation; partnership with government, government agencies, professional societies or charitable organizations to plan and/or implement change; other institutional, regional or system-level changes or improvements, etc.

- Societal

Social, cultural, environmental, and economic returns from research outputs/products and associated outcomes [some papers seem to put patents, companies, adoption by government, etc. here; we can clarify concurrent with reviewing studies/documents on social impact of research]

- Purpose of assessment includes hiring, annual review, re-appointment, compensation, bonuses, tenure, promotion, consideration for leadership roles or awards, etc.
- Policies or approaches or practices related to implementing or applying measures of research productivity; for example, training appointments committee members in DORA principles, strategies for creating culture change, etc.

**Publication design/type** [studies/documents eligible for review may include the following types/formats]

- Published research identified in indexed databases:
  - Studies that employed quantitative, qualitative, mixed/multiple methods or expert consensus to generate, compile, apply or evaluate research productivity principles, processes, measures, etc.
  - Reviews (e.g. scoping, systematic) of published research are not eligible but we will screen references for eligible primary studies

**Outcomes** [studies/documents eligible for review may assess/report one or more of the following]

- Criteria, policies or practices for implementing/applying criteria, or related recommendations
- Knowledge, attitudes, experiences or preferences of researchers or research institutes pertaining to criteria or related policies or practices to implement/apply them; or actual or anticipated enablers/barriers
- Actual or potential beneficial and unintended/unanticipated outcomes/impacts or implications of implementing/applying criteria or policies/practices

**NOT ELIGIBLE**

- Studies in which participants are largely students, trainees, fellows, research assistants/coordinators/associates
- Identification or appraisal of competencies required to practice a particular profession/required of trainees
- High level principles on the need for measures other than JIF (i.e. aspirational statements similar to DORA)
- Evaluation of research productivity by research funding organizations or panels in the context of funding competitions (rather than institutional personnel decisions); or the impact of funding/awards on research productivity (measured in terms of research money or publications)
- Metrics for assessing research performance of organizations (e.g. total research funding or a university department or research institute), or health systems/countries (e.g. identifying impacts reported in research funding databases)
- Approaches or interventions for faculty development or continuing professional development (e.g. mentoring) to help them become productive researchers
- Research framed as return on investment in non-health care sectors (e.g. agriculture, construction industry)
- Assessment of performance in areas other than research (e.g. clinical income, service to institution or external organizations, supervision or mentoring of trainees or staff, teaching, clinical quality improvement, etc.)
- Focuses on alternative ways of assessing the number or quality of publications, or co-publishing author networks
- Development or validity of different ways to calculate journal impact factor or other publication metrics
- Focus on number/value of research grants (alone, or in addition to publication metrics)
- Authorship or peer review practices: refers to studies of the rules imposed by journals or research organizations on number/order of authors, author contributions, peer review procedures, etc.
- Assessment of or strategies/interventions to promote inclusivity, diversity, equity or accessibility (also germane to hiring/promotion and other decisions, but out of scope for this review)
- Studies or documents that recommend against using JIF, or recommend using other criteria (outputs, measures, impact, etc.) to assess research productivity, but do not specify alternatives
- Publication type: editorials, letters, commentaries that address previous bullet point
