## Supplemental Appendix 2 for "DORA-compliant measures to assess research quality and impact in biomedical institutions: review of published research, international best practice and Delphi survey"

**APPENDIX 2. MEDLINE search strategy**

| # | Search Statement | Results |
| --- | --- | --- |
| 1 | Personnel Selection/ | 12993 |
| 2 | personnel management/ or employee incentive plans/ or employee performance appraisal/ or academic performance/ | 20724 |
| 3 | career mobility/ | 11623 |
| 4 | Research Personnel/ | 16831 |
| 5 | faculty/ or faculty, medical/ or faculty, nursing/ | 33395 |
| 6 | biomedical research/ or exp genetic research/ or exp health services research/ or human experimentation/ or exp nursing research/ or exp outcome assessment, health care/ or exp pharmacy research/ or exp rehabilitation research/ or exp stem cell research/ or exp translational medical research/ | 1466055 |
| 7 | exp Behavioral Sciences/ | 252249 |
| 8 | cognitive science/ or cognitive neuroscience/ | 1267 |
| 9 | Biomedical Engineering/ | 11126 |
| 10 | Biostatistics/ | 2214 |
| 11 | Nutritional Sciences/ | 11765 |
| 12 | or/1-3 | 42278 |
| 13 | or/4-11 | 1765462 |
| 14 | 12 and 13 | 6799 |
| 15 | guideline/ | 16343 |
| 16 | benchmarking/ | 13780 |
| 17 | indicator*.mp. | 378437 |
| 18 | standard*.mp. | 1962023 |
| 19 | [productivity.mp](http://productivity.mp/). | 62650 |
| 20 | achieve*.mp. | 988610 |
| 21 | accomplish*.mp. | 101486 |
| 22 | altimetrics.mp. | 1 |
| 23 | (performance adj2 measure*).mp. | 23572 |
| 24 | or/15-22 | 3301091 |
| 54 | 14 and 23 | 2215 |
| 25 | limit 24 to (english language and humans and yr="2013 -Current") | 380 |
| 26 | limit 25 to (comment or editorial or interview or lecture or letter or news) | 42 |
| 27 | 25 not 26 | 338 |
