## Supplemental Appendix 3 for "DORA-compliant measures to assess research quality and impact in biomedical institutions: review of published research, international best practice and Delphi survey"

**APPENDIX 3. Data extracted from articles included in the scoping review**

| Study | Objective | Design/Participants | Results |
| --- | --- | --- | --- |
| Aquaviva 2020 [28]  United States  Health professions | Develop a method for scholars in the health professions to cite social media activities and impact | Joint development of a guideline via Twitter  39 respondents; characteristics not reported | Overall Reach (Time Period: x/x/xx to y/y/xx)  [Platform]: [username]  Number of Followers/Subscribers/Connections:  Number of [Tweets, Posts, Videos, etc]:  Total Impressions and/or Other Platform-Specific Metrics:  Select Social Media Contributions  Innovation (contributions that propose new ideas)       Link to contributions:       Number of impressions:       Explanation of why the scholar chose to highlight this:  Dissemination (contributions that share resources and/or findings)       Link to contributions:       Number of impressions:       Explanation of why the scholar chose to highlight this:  Education (contributions that teach people something)       Link to contribution:       Number of impressions:       Explanation of why the scholar chose to highlight this:  Advocacy (contributions about changing laws, policies, practices, and/or systems)       Link to contribution:       Number of impressions:       Explanation of why the scholar chose to highlight this:  Mentorship (contributions about mentees achievements)       Link to contribution:       Number of impressions:       Explanation of why the scholar chose to highlight this contribution:       Presentations/Chats/Blogs/Podcasts Delivered Via Social Media  [Platform] Chats       Name of [Platform] Chat:       Role: [host, cohost, etc]       If longitudinal, list time period and cite metrics. If discrete, list dates and topics:       Explanation of why the scholar chose to highlight this contribution:  [Platform] Live Video       Host or cohost(s) of live feed:       Individual sessions [Date(s), Topic(s), Engagement/Reach]:       Permanent Link:       Explanation of why the scholar chose to highlight this contribution:  [Platform] Recorded Video       Host or cohost(s) of recorded video:       Title of Video:       Date(s), Topic(s), Engagement/Reach:       Permanent Link:       Explanation of why the scholar chose to highlight this contribution:  Blog Posts       Author:       Title of Blog:       Organization/Entity Publishing the Blog:       Date Published:       Link to Blog:       Explanation of why the scholar chose to highlight this contribution:  Podcasts       Role: [Host or Guest]       Name of Podcast:       Date Released:       Link to Podcast Episode:       Explanation of why the scholar chose to highlight this contribution:       Infographics/Other Visuals  Infographic       Title of Infographic:       Date Posted/Published:       Link to Infographic:       Explanation of why the scholar chose to highlight this contribution:  Other Visuals       Author/Creator:       Title of Visual:       Date Posted/Published:       Link to Visual:       Explanation of why the scholar chose to highlight this contribution: |
| Clement 2020 [29]  United States  Biomedical sciences | Explore qualifications required to obtain a faculty position in biomedical life sciences | Multiple methods: interviews with 23 biomedical life science faculty followed by survey in which 19 participated  58% women; 26% visible minority | - Clear and compelling verbal communication of research - First author publications - Interesting research vision with feasible and appropriate methods - Awareness of funding organizations and plan for acquiring funding - Evidence of research independence - Letters of recommendation that emphasize candidate’s potential to be a leader in the field - Demonstrated collegiality/collaboration - Fit/synergy of research interests/plan with organizational priorities |
| Husain 2020 [30]  United States  medicine | To generate consensus on how to evaluate the impact of digital scholarly work for promotion and tenure | Nominal group consensus process involving X at a meeting of the Council of  Emergency Medicine Residency Directors  Number and characteristics not reported | IMPACT (shows work reached intended audience)  Pageviews, Time Spent on Page, Likes, Impressions, Dissemination (Shares), Unique Users, Geographic Reach, Followers on Professional Social Media Accounts, Social Media Index, Digital Object Identifier (DOI), Alexa Ranking, Altmetrics  Example: Pageviews 4137; Altmetric Score 61; 202 tweets from 86 users, with an upper bound of 263,362  Followers  ROLE (demonstrate “brand” to establish area of expertise)  Editor, Author, Curator, Reviewer, Invited Commentaries, Podcast Guest or Editor  Example:  [Invited Commentary] Berg A, Weston V, Gisondi MA. Journal Club: Coronary CT Angiography Versus Traditional Care. NUEM Blog. http://www.nuemblog.com/blog/cta-for-chest-pain/ Published online 4/12/16.  QUALITY (highlight novel quality assurance methods unique to digital scholarship)  METRIQ-5 and -8, rMETRIQ, ALiEM AIR Score, SAEM Online Academic Resources, (SOAR)  Social Media Index (SMi), The Quality Checklists for Health, Professions Blogs and Podcasts  Example:  [Peer-reviewed blog] Long, B. “Myths in Heart Failure: Part I – ED Evaluation” emDOCs.net http://www.emdocs.net/myths-in-heartfailure-part-i-ed-evaluation/ published online 7/23/2018.  Selected as ALIEM AIR Cardiovascular, Non-ACS module 2019. This post was deemed to be of an acceptable score within the ALiEM AIR Scoring tool, and was granted the designation “AIR  Approved” by the adjudicating group of educators. There is a second tier below, known as “honorable mention” for posts of moderate quality that did not meet the threshold for inclusion. |
| Rice 2020 [31]  Canada  Biomedical sciences | Assess criteria used in promotion and tenure decisions by international faculties of biomedical sciences | Content analysis of guidelines at 92 institutions among 146 with a faculty of biomedical sciences | - Traditional criteria of peer reviewed publications, authorship order, journal impact factor, grant funding, and national or international reputation were mentioned in 95% (n=87), 37% (34), 28% (26), 67% (62), and 48% (44) of the guidelines, respectively. - Mention of alternative metrics for sharing research (3%; n=3) and data sharing (1%; 1) was rare, and three criteria (publishing in open access mediums, registering research, and adhering to reporting guidelines) were not found in any guidelines reviewed |
| Aizer Brody 2019 [32]  United States  Nursing | Explore whether interdisciplinary team science was recognized in nursing school appointment, promotion and tenure (APT) criteria | Content analysis of 18 APT criteria documents at 18 research-intensive schools of nursing | - 8 of 18 documents included any reference to team science principles, but these mentions were indirect and brief, and included leading a collaboration, or being a member of a collaboration - 5 of 18 documents identified specific activities:   - Lead national committees/organizations   - Receive awards   - Deliver invited lectures   - Mentoring |
| Klein 2019 [33]  United States  Pharmacy | Assess criteria for promotion used by research-intensive pharmacy programs | Content analysis of promotion guidelines at 10 programs | - Criteria varied across programs - Criteria relevant to research included:   - Publications – abstracts, book chapters, original research, review articles, author order, collaboration, impact factor, independent work   - Grants – number of applications, number of funded grants, funding source   - Recognition – invited presentations, innovative practices, honours/awards, committee membership, leadership roles, journal reviewer |
| LeMaire 2018 [34]  United States  Surgery | To develop a scoring system for academic productivity linked to faculty compensation | Before-after cohort study of the impact of a self-reporting scoring-compensation system on productivity  319 faculty, characteristics not reported | Self-report measures of research productivity included presentations, publications, submitted grants, funded grants, industry-sponsored trials, committee leadership and scientific manuscript peer review  The bonus system resulted in significant increases in:   - presentations (579 to 862; P ¼ 0.02; 49% increase) - publications (390 to 446; P ¼ 0.02; 14%) - total research funding ($4.6M to $8.4 M; P < 0.001; 83%) - NIH funding ($0.6M to $3.4 M; P < 0.001; 467%) - industry-sponsored clinical trials (8 to 23; P ¼ 0.002; 188%), - academic society committee positions (226 to 298; P < 0.001; 32%) - editorial leadership positions (50 to 74; P ¼ 0.01; 48%) |
| Moher 2018 [35]  Canada  Clinical and life sciences | To generate principles to assess scientists for hiring, promotion and tenure | Consensus meeting of 22 international academic leaders, funders, and scientists who reviewed criteria extracted from a literature review, and proposed solutions  8 women | - Address societal needs (need for research to establish how to do this) - Contributions to science – in addition to more traditional measures, also assess registration, sharing, contributions to peer review and alternative metrics - Transparent publishing of all research regardless of results – if not through formal publication, use preprints or OA repositories - Reward open research – sharing of data, protocols, software, code, materials and other research tools - Complementary recommendations: fund more research to generate evidence that informs the assessment of science and faculty; promote funding of grant applications without specific aims that pursue a broad investigative agenda |
| Sehgal 2017 [36]  United States  Medicine | Evaluate the impact of adopting a quality improvement portfolio on career advancement | Cohort study in the faculty of medicine at one university | - 67 QI portfolios were submitted (61% women) - 100% of faculty received their requested academic advancement - 83% agreed it was an effective tool for recognizing faculty contributions in QI work |
| Finney 2016 [37]  United States  Health services research | Assess academic productivity of recipients of career development awards from three national programs | Content analysis of CV’s of 242 career award recipients: 50% women, 20.4% minority groups | Measures used to assess productivity:   - Tenure - Number of grants as primary investigator (and number over $100K) - Journal articles as first author - h-index score - membership in major granting review panel - mentoring postgraduate researchers |
| Hicks  2015 [14]  United States  Biomedical sciences | Ten principles to guide research evaluation | Distillation of best practice in metrics-based research assessment that were generated at a conference by invited experts | 10 Principles of the Leiden Manifesto:   - Quantitative evaluation should support qualitative, expert assessment - Measure performance against the research missions of the institution, group or researcher - Protect excellence in locally relevant research - Keep data collection and analytical processes open, transparent and simple - Allow those evaluated to verify data and analysis - Account for variation by field in publication and citation practices - Base assessment of individual researchers on a qualitative judgement of their portfolio - Avoid misplaced concreteness and false precision (uses multiple indicators to provide a more robust and pluralistic picture) - Recognize the systemic effects of assessment and indicators (a suite of indicators is always preferable — a single one will invite gaming and goal displacement) - Scrutinize indicators regularly and update them |
| Kairouz 2014 [37]  United States  Medicine | To assess how faculties of medicine measure productivity for the purpose of salary compensation | Survey of Chairs of academic Departments of Medicine: 78 of 152 responded | - Productivity assessed based on clinical (98%), research (61%), teaching (62%) and administrative (64%) activities - Departments reported a wide variation of what exact activities are measured - For research, this included peer-reviewed publications, career awards, research grants awarded and external recognition |
