## Supplemental Appendix 4 for "DORA-compliant measures to assess research quality and impact in biomedical institutions: review of published research, international best practice and Delphi survey"

**APPENDIX 4. Data extracted from grey literature review documents**

**Canadian Universities**

| Document | Organization type | Purpose | Indicators/Measures | Strategies |
| --- | --- | --- | --- | --- |
| Faculty of Medicine, University of Calgary  Criteria for appointment, promotion, merit increment and tenure of full-time faculty [38]  December 2008 (17 pages) | Academic institution | Criteria for evaluation of research output | General  Productivity and quality should rise as faculty move through the academic ranks. A junior faculty member might be expected to develop skills in independent research by demonstrating the capability to initiate and  maintain innovative research protocols and programs. More senior investigators may receive special merit for their ability to recruit promising students and junior colleagues, and by providing for them an atmosphere which is conducive to the development of distinguished careers in research.  First order criteria:   - Peer-reviewed publications (quality of the publication, journal reputation, nature of authorship) - Acquisition of refereed grants - Invited presentations at symposia, national and international meetings - Leadership roles in fostering research (e.g. coordination of multidisciplinary collaborative group grants or equivalent, coordination of conferences or symposia, chair of national or provincial society of research scientists   Second order criteria   - Commercialization of technology (patents, licenses) - Non-peer reviewed publications (e.g. book chapters) - Local, national and international communications (e.g. oral or poster presentations at meetings) - Non-refereed grants or contracts - Participation in networks or consortia (e.g. interdisciplinary research teams) | --- |
| University of Victoria,  Faculty of Human and Social Development  Faculty Evaluation Policy 2019-2022 [39]  January 2020 (56 pages) | Academic institution | Criteria and procedures for the evaluation of faculty members for the purposes of reappointment, tenure, promotion, continuing appointment, and salary adjustment on the basis of demonstrated achievement. | - Peer-reviewed papers and scholarly papers - Other forms of creative achievement relevant to discipline - Research grants, awards and fellowships - Activities and outputs related to community engaged research partnerships (co-production of research with those outside of academia) - Collaborative and interdisciplinary scholarship - Appointments to professional and scholarly adjudicatory or review boards our councils - Recognition by learned professional societies - Evidence of reputation for scholarship - Number of research grant applications (not just those that are successful)   Examples of scholarship include but are not limited to:   - peer-reviewed publications (see below); - non-peer-reviewed material (see below); - films, videos, computer software, web-sites, pod-casts; - tests, questionnaires, or assessment instruments; - research grants and contracts (see below); - research proposals; - conference presentations; - invited addresses to professional associations/societies/community groups; - editing a research or professional journal; - invited contributions to policy development - developing a new practice technique; - building university-community partnerships; - developing research protocols; - distance education, distributed or blended learning course development; and - artistic creations and productions. | Every Unit must develop examples and indicators in their Standard that are consistent with each stage of career to identify Research which meets performance expectations and Research which exceeds performance expectations  To assess research on its own merits rather than on the journal in which it was published….faculty are encouraged to provide evidence of the quality, significance and impact of their scholarship (refereed and non-refereed, written and oral) and so may include assessments from a range of users (e.g. academic peers, government officials, NGOs officers, Indigenous community leaders, conference participants, academic and community awards, community members, patients, youth).  They should also specify the level of impact: individual, community, system, population |
| University of Regina, Faculty of Science  Criteria document for faculty members and instructors. Terms of reference for assignment of duties, performance review, career progress, and sabbaticals [40]  March 2017 (26 pages) | Academic institution | To provide criteria by which recommendations and decisions are made on duties, performance review, merit, tenure, promotion and sabbatical | Scholarship involves the contribution of new knowledge (i.e., research) and scholarly dissemination of knowledge (e.g., refereed articles, books, reviews) in the respective disciplines.  The publication of research results in books and papers, or presentations at conferences are easily documented. Evidence of prestige among colleagues in the international scientific community is also a useful yardstick of research ability. Invitations to present papers, chair conference sessions, participate in symposia, or referee papers and research grant applications, provide supporting evidence of scholarly recognition.  A simple counting of publications without assessment of their quality is not sufficient, and indeed may be misleading. Consequently, care must be exercised in evaluating publications. Peer evaluation of such work is critical and faculty members must publish in peer-evaluated media acceptable to their fields. | The Review Committee consists of one regular member and one proxy member from each department, elected for two-year terms. Faculty members may serve multiple terms as regular members of the Faculty Review Committee; however, no faculty member shall serve two consecutive terms in this capacity. |
| University of Alberta, Faculty of Medicine & Dentistry  Procedures and criteria for tenure, promotion, merit and sabbaticals [41]  November 2017 (25 pages) | Academic institution | To describe procedures and criteria for performance review | Tenure  Documentation of how their major job domain has had an impact at least at a local level, or preferably to an emerging/established national recognition  Full Professor  Established local and national, and emerging international recognition in that domain.  Performance Standards:  Excellent  Performance that is: a) functioning beyond commendable for their rank and/or percentage of position description, and/or b) distinguishing and expanding skills/learning opportunities/stewarding our people, our work and service  Commendable  Performance that is beyond Acceptable will be distinguished for merit  Acceptable  Performance demonstrates a significant deficiency in one domain of evaluation, but performs well in the other domains, or when the faculty member overall performs below expected for rank, but remains within the acceptable range  Unacceptable  Academic performance is unsatisfactory and unacceptable  Criteria   - Any activity that is peer-reviewed, disseminated and has some measure/description of its impact and outcome is scholarly output (scholarship/research). - Peer reviewed, competitive, external (e.g. tri-council) funding as PI - Unsuccessful funding applications may be considered for merit depending on the quality/competitiveness of the funding, and the rank and percentage research time of the faculty member. - Faculty are expected to share knowledge by making it public, whether via publication, correspondence, presentations or pedagogy. New technologies make such exchange even more widely possible than ever before. - Presentations at various venues: conferences, academic institutions, academic meetings are of higher quality than presentations to the media, lay-community; depending on the field, an oral presentation to a large international audience may have more impact than a publication. - Social Media: blogs, Twitter, webpage, YouTube, etc. are lower quality venues, unless peer-review, uptake and impact can be described. - Public Compendiums: MedEDPortal, MedEDPublish, vodcasts, podcasts. - Publications (print or electronic): books, book chapters, original research articles, review articles, guidelines, case reports/series, editorial; an abstract as part of a conference proceeding should be listed as a product of scholarship or with presentation.   FEC expects faculty members with a lower percentage research (10%) and rank (Assistant Professor) are expected to have at least one product or process of scholarship or item of dissemination per reporting year. Faculty members with a higher percentage research and/or higher academic rank, are expected to have more items and/or of increasing quality (broader scope, greater impact) of scholarship.  Many research activities are collaborative; for each category (funding, products and processes, and dissemination), the faculty member should describe their role and contributions. As well, they should describe the scope (local, national, international, etc.) of their work. To have impact, and if known, faculty members should indicate how the research had a reported change to target group (e.g. patients, learners, faculty colleagues, government, communities), and for influence, how the work has been used by other groups.  The FEC considers not just the quantity of scholarly products, but also their quality, which for traditional discovery scholarship offers publication opportunities with reported impact factors and citation indices. Other forms of valued scholarship and innovation (e.g. education scholarship, community engagement, creative academic work, entrepreneurship) presently do not have similar metrics. As such, faculty members engaged in these activities should describe their role, scope and importantly the use and impact of their work as fully as possible.  Partnerships with industry are a component of scholarly work of many FoMD faculty members. Funding or dissemination that is sponsored by industry may be viewed as less rigorous than other competitive, peer-reviewed sources.  All scholarly endeavors are time consuming activities; some, especially those of higher quality, need development for more than one or more academic reporting years. During this time, funding, productivity and/or dissemination may appear low. A thorough annual report that describes the potential impact and quality of work may assist the Department Chair and FEC in such cases. However, for FEC to assess work without any tangible product of scholarship, the faculty member must describe the process of scholarship and demonstrate appropriate progress. | To facilitate success, each Department Chair shall:   - Review with each member their responsibilities and expectations - Meet with each member annually to discuss their annual report; and performance quality, progress and trajectory - Discuss career goals, and offer mentorship and other supports - Discuss merit recommendations   Annually, the performance of each member will be reviewed by the Department Chair and Faculty Evaluation Committee according to the member’s position description  Probationary periods: first period = 4 years, second period = 2 more years, but could be extended by the Faculty Evaluation Committee depending on circumstances  To help members plan research trajectories and prepare for review, the Department should provide a manual that describes the process, and providers examples of lower and higher quality research activities for each discipline |

**International Organizations**

| Document | Organization type | Purpose | Indicators/Measures | Strategies |
| --- | --- | --- | --- | --- |
| Declaration on Research Assessment (DORA) [1]  <https://sfdora.org/>  Viewed March 2021 | Advocate | To advance practical and robust approaches to research assessment globally | --- | Rethinking Research Assessment: Ideas for Action  (infographic to help organizations understand barriers to change)   - Instill standards and structure into research assessment processes - Foster a sense of personal accountability in faculty and staff - Prioritize equity and transparency of research assessment processes - Take a big picture or portfolio view toward researcher contributions - Refine research assessment processes through iterative feedback   Rethinking Research Assessment: Unintended cognitive and systems biases  (infographic: 7 personal biases that can influence hiring, promotion and tenure decisions; strategies to develop institutional conditions that reduce bias)   - Recognize where old assumptions may overly reward those who are more traditionally successful, at the expense of new or more diverse talent - Set, publicize, and adhere to measurable goals that look beyond traditional norms of success when reviewing potential candidates to broaden the pool of individuals under consideration - Use structured interview protocols to keep decision-makers focused on agreed-upon qualities, rather than on reputation - Explicitly articulate and consider long-term and qualitative values, as well as short-term or easily quantifiable needs - Have applicants highlight and articulate their most meaningful contributions to reduce reviewer reliance on journal names or quantifiable characteristics of productivity - Balance the use of quantitative metrics with qualitative inputs, like narrative CVs, that capture more intangible qualities - Select standards based on a wide set of inputs rather than a narrow or anecdotal set - Recognize where setting specific, quantifiable goals may be reinforcing some behaviors at the expense of others - Assemble diverse teams—across gender, seniority, cultures, and under-represented minoritized populations—to bring a range of perspectives and experiences into decisions - Look outside your institution or discipline to broaden a sense of “normal” - Put reputation-based indicators like education at the end of applicant materials to reduce preconceived notions |
| Wilsdon et al., Chair, University of Sussex with an international steering committee  The Metric Tide. Report of the independent review of the role of metrics in research assessment and management [42]  July 2015 (178 pages) | Special interest | To ensure that indicators and underlying data infrastructure support the diverse qualities and impacts of research  Based on literature review and stakeholder consultation in focus groups and workshops | Metrics should support, not supplant, expert judgement through peer review  Carefully selected and applied quantitative indicators can be a useful complement to other forms of evaluation and decision-making  One size is unlikely to fit all: a mature research system needs a variable geometry of expert judgement, quantitative and qualitative indicators.  Research assessment needs to be undertaken with due regard for context and disciplinary diversity.  Quantitative indicators can be gamed, or can lead to unintended consequences; journal impact factors and citation counts are two prominent examples. These consequences need to be identified, acknowledged and addressed. | Leadership/Governance   1. The research community should develop a more sophisticated and nuanced approach to the contribution and limitations of quantitative indicators 2. Institutional leaders should develop a clear statement of principles on their approach to research management and assessment 3. Research managers and administrators should champion these principles and the use of responsible metrics within their institutions 4. HR managers and recruitment or promotion panels should be explicit about the criteria used for academic appointment and promotion decisions 5. Individual researchers should be mindful of the limitations of particular indicators in the way they present their own CVs and evaluate the work of colleagues 6. Research funders should develop their own context-specific principles for the use of quantitative indicators in research assessment and management and ensure that these are well communicated, easy to locate and understand 7. Data providers and analysts should strive for greater transparency and interoperability between different measurement systems 8. Publishers should reduce emphasis on journal impact factors as a promotional tool   Data Infrastructure   9. A set of principles should be developed for technologies, practices and cultures that can support open, trustworthy research information management 10. The research system should take full advantage of ORCID as its preferred system of unique identifiers. ORCID iDs should be mandatory for all researchers 11. Identifiers are also needed for institutions, and the most likely candidate for a global solution is the ISNI 12. Publishers should mandate ORCID iDs and ISNIs and funder grant references for article submission 13. The use of digital object identifiers (DOIs) should be extended to cover all research outputs 14. Further investment in research information infrastructure is required   Enhancing Existing Data Sources   15. Explore how to leverage data held in existing platforms to support research assessment 16. Identify ways of linking data gathered from research-related platforms   Using Metrics   17. In assessing outputs, quantitative data – particularly around published outputs – should continue to have a place in informing peer review judgements of research quality; in assessing impact, develop clear guidelines for the use of quantitative indicators; in assessing the research environment, consider quantitative data with sufficient context to enable their interpretation   Coordinating Activity   18. Establish a multi-level forum to work on issues of data standards, interoperability, openness and transparency 19. Research funders need to increase investment in the science of science policy 20. Contribute to discussion at [www.ResponsibleMetrics.org](http://www.ResponsibleMetrics.org) |
| Metrics Toolkit  Based at IUPUI, Indiana; supported by Altimetrics (Wiley, Taylor & Frances) [43]  <https://www.metrics-toolkit.org/>  Viewed March 16, 2021 | Special interest | Online resource that provides information about research metrics across scholarly disciplines to help educate individuals in the academic community. It gives a summary of how each metric is calculated, what it can be applied to, what it’s limitations are, and lists appropriate and inappropriate use cases | Altimetrics Attention Score  Amazon Ratings and Reviews  Blog mentions  Citations - articles, citation percentiles, highly cited labels  Citations - books and book chapters  Citations – data or software  Citation – field normalized impact  Downloads – articles, book chapters, software, etc.  Facebook – posts, comments, likes and shares  Github – forks, collaborators, watchers  Goodreads – ratings and reviews  H-index  Journal acceptance rate  Journal impact factor  Mendeley Readers  Monograph holdings  Monograph sales and ranking  News mentions  Policy mentions  Publons score  Pubpeer comments  Relative citation ratio  Twitter mentions  Wikipedia citations | --- |
| International Network of Research Management Societies (INORMS) Research Evaluation Working Group [44]  <https://inorms.net/>  Viewed March 16, 2021 | Special interest | Principles for responsible research evaluation and a process to implement good evaluations | START with what you value  ➢ Not with what others’ value (external drivers)  ➢ Not with available data sources (the ‘Streetlight Effect’)  CONTEXT considerations  ➢ WHO are you evaluating? (Entity size)  ➢ WHY are you evaluating?  OPTIONS for evaluating  ➢ Consider both quantitative and qualitative options  ➢ Be careful when using quantities to indicate qualities  ➢ Evaluate with the evaluated  PROBE deeply  ➢ WHO might your evaluation approach discriminate against?  ➢ HOW might your evaluation approach be gamed?  ➢ WHAT might the unintended consequences be?  ➢ Does the cost outweigh the benefit?  EVALUATE your evaluation  ➢ Did your evaluation achieve its aims?  ➢ Was it formative as well as summative?  ➢ Keep your approach under review | Maintain institutional autonomy  ➢ INFORMAL: Control your own destiny  ➢ External evaluations (e.g., University Rankings) are a fact of life but mission- and value-led organisations should consider carefully what those evaluations actually assess, and not seek to improve their performance against measures that are not aligned to their organisational mission or values.  ➢ Campbell’s Law tells us that we get what we measure. Institutions need to measure what matters to them, in order to generate outcomes that are aligned with their mission.  Make better decisions  ➢ INFORMAL: Play stupid games win stupid prizes  ➢ Evaluations that use indicators that are an inappropriate proxy for the things they seek to measure will lead to poor decision-making and unintended consequences.  ➢ A combination of quantitative and qualitative (peer review) approaches usually provide better answers to our evaluation questions.  ➢ There are many well-documented negative systemic effects of poor evaluation approaches, including short-termism, goal displacement, and discouraging initiative and innovation.  Ensure return on investment  ➢ INFORMAL: Getting your money’s worth  ➢ Research evaluation costs money and institutions should weigh up the cost-benefit ratio of undertaking any assessment. It may be that their ends can be achieved another way.  ➢ Institutions need to ensure they are investing wisely in meaningful evaluations that are actually going to give them valid answers to their questions, and not waste money on short-cuts that won’t.  Establish operational readiness  ➢ INFORMAL: Tick all the right boxes  ➢ There is an increased focus on responsible research evaluation as a result of sector agendas including open research, improving research culture and responsible research and innovation.  ➢ Responsible research evaluation is now a requirement of some research funder open access policies.  ➢ The Global Research Council is taking an interest in this area and further funder expectations may follow.  Manage reputational risk and enhance staff well-being  ➢ INFORMAL: Avoid bad press  ➢ As a result of sector agendas, the misuse of research metrics is coming under increased scrutiny.  ➢ The over-use of poor metrics has a negative impact on staff well-being and mental health.  ➢ There have been some high-profile cases where poor evaluations have led to tragic consequences. |
| International Development Research Centre (IDRC), Canada  RQ+ Research Quality Plus. A holistic approach to evaluating research (2016) [45]]  <https://sfdora.org/wp-content/uploads/2020/11/IDL-56528.pdf>  Viewed March 16, 2021 | Special interest | International collaboration to develop a new approach to evaluating the quality of  research | Key Influences   - Maturity of the research field - Research capacity strengthening - Risk in the research environment - Risk in the political environment - Risk in the data environment   Research Quality Dimensions   - Research integrity - Research legitimacy - Research importance - Positioning for use   Evaluative Rubrics  Provides criteria by which to judge each of the above quality dimensions not applicable, unacceptable, less than acceptable, acceptable to good, and very good | --- |
| Royal Society  (based on London UK, society of international scientists)  Research Culture: Changing Expectations. Conference Report [46] | Special interest | To reflect on and generate a vision of a more robust research enterprise | Résumé for Researchers (template to report research achievements)  How have you contributed to the generation of knowledge?  ideas, hypotheses, skills, dissemination, funding, awards, and outputs: open data sets, software, publications, commercial, entrepreneurial or industrial products, clinical practice developments, educational products, policy publications, evidence synthesis pieces and conference publications | Recommendations:  1/  Need to create a more porous research culture, and enable more mobile and flexible research careers. Moving towards a structure for research careers that involves multiple, intersecting, pathways, and that allows researchers to work in and develop knowledge of other sectors and practices should be a key ambition for us all.  2/  Redefine or broaden our notion of what  ‘research leadership’ is. Moving to a concept of inclusive and collaborative leadership, which recognises a responsibility to and reciprocity with colleagues  3/  We need to change the academic CV, so that people are judged on what they have done broadly in academia, their advances in the field, their links with industry and society, and their contributions to knowledge exchange and outreach. Evidence for this isn’t just in the form of papers and books, but might be datasets, online teaching resources, a television series, or a patent. This means that the CV can be shaped, naturally, for different subject areas and disciplines as well as cross-discipline research. |
| Canadian Academy of Health Sciences  Making an impact: A preferred framework and indicators to measure returns on investment in health research [47]  January 2009 (136 pages) | Special interest group | To define the impacts of health research | SEE details of numerous multi-level indicators on pages 25-32  **many not recommended to assess individual researchers (more suitable for organization or system level) | --- |
| European University Association  Reimagining Academic Career Assessment: Stories of innovation and change [48]  January 2021 (47 pages) | Academic | Includes 9 university case studies in responsible academic  career assessment selected from DORA signatories and SPARC Europe and EUA members |  | Overall case study findings:   - Move away from quantitative publication metrics to a more holistic approach that rewards a broader range of academic activities - Aiming to improve the academic culture at their institution long-term - All cases highlight an absence of knowledge about how to incentivize and reward a broader range of academic activities (i.e. information that would be used in evaluations in lieu of quantitative publications) - More accurate, transparent, and responsible approaches to academic evaluation should not primarily or even necessarily aim to add more indicators, but rather seek to find dynamic, context-sensitive, and above all holistic approaches that allow researchers and universities the freedom to pursue/manage academic activities in any way they believe is most effective in service to society - To this end, institutions should also be flexible and provide opportunities to revisit and refine policies and practices as needed. - Raising awareness, community engagement, and building capacity are key to expand the range of academic activities that are being incentivized and rewarded |
| University of Catalunya, Spain  Open Knowledge Action Plan: Frame of action [49]  May 2019 (27 pages) | Academic |  | Six areas of focus:  Open access publications  % of articles published in open access journals  Includes articles, books, book chapters, etc.  Open FAIR data  All data/knowledge created follows FAIR principles: Findability (meta-data DOI), Accessibility (open protocol and data), Interoperability (data use widely-applied language/format), Reuse of digital assets (data are richly described)  Open learning  Learning resources are fully rather than selectively open, with copyright exceptions  Open innovation  Research conducted via open platforms that enable local, national and international collaboration to generate solutions to societal challenges  Open to society  To be integrated with society through forums that that enable co-creation of knowledge based on social needs to generate knowledge with high social impact  Research evaluation models  Transition to a more qualitative evaluation that incorporates constant learning and transformation | Create awareness among the research community of the relatively novel concepts of open knowledge  Make initiatives visible so that they can serve as examples to follow  All communication pertaining to research must be made consistent with the principles of open knowledge  Technological infrastructure is needed to enable the availability of open knowledge platforms  Create an action plan to implement open knowledge changes  Regularly monitor implementation and progress of the action plan |
| Ghent University, Belgium  Vision Statement for Evaluating Research  November 2016 [50]  Portfolio of Research Dimensions  No date (10 pages) | Academic | To describe goals and measures for research assessment | From Portfolio of Research Dimensions:  VITALITY   - Favourable evaluation of applications for research funds from external funding agencies (FWO (Research Foundation Flanders), Europe, foundations, industry) - International awards and recognitions based on intrinsic research quality, such as ERC Advanced Grant, Francqui Chair, etc. and general or field-specific special recognitions - Reviews of papers as a peer review expert - Leading role as an evaluator or expert in the researcher’s field (e.g. panel chair, member of a “high level group”) - Sustained, outstanding track record in (social) entrepreneurship - Pioneering role in a spin-off process - Pioneering role in a scientific organisation - Publications with an important personal contribution (in peer-reviewed journals, a book publication, peer-reviewed conference proceedings, …) - Keynote speeches (by invitation) at prestigious conferences - Growth of the research group (in FTE, for example) - Successful doctoral defences as supervisor - Review articles by invitation in renowned specialist journals   ORIGINALITY   - Development of a new research line within the group - Recognition within the international research community, such as with an ERC grant or other general or field-specific special recognitions - International, top-level awards, both general and field-specific - Appreciation from evaluators/excellent score for originality in applications for research funds from external funding agencies (FWO (Research Foundation Flanders), Europe, foundations, industry) - Development and testing of new methodologies - Negative review in a leading journal due to an approach that is too radical or innovative - Normalised citation impact of SSCI and SCIE publications during the reference period - Publications in top 5% or 10% journals - Book publication by an internationally-renowned publisher - Published negative results - Successful doctoral defences as supervisor   LEADERSHIP   - Role as a people manager in the design of a research group or research consortium, - Supervision of employees - Appropriate management and/or restructuring of the research group - High-quality supervision of doctoral researchers - Growth of the research group (in FTE, for example) - Leading role as an evaluator or expert in the researcher’s field (e.g. panel chair, member of a “high level group”) - Reviews of papers as a peer review expert - Pioneering role in a spin-off process - Pioneering role in a scientific organisation - International awards and recognitions for high-level research, such as ERC advanced grant, Francqui Chair, etc. and general or field-specific recognitions - Keynote speeches (by invitation) at prestigious conferences - Successful doctoral defences as supervisor - Research budget obtained in the previous 5 years   INTERDISCIPLINARITY   - Sharing of research infrastructure - Management of and involvement in an interdisciplinary research group - Detailed strategy for the interdisciplinary approach in the researcher’s own research environment - Implementation within the researcher’s own research group of an “Open Science” culture towards other fields - Contribution to interdisciplinary journals or specialist journals - Joint publications with colleagues in other fields - Joint doctoral projects with colleagues in other fields (including Ghent University interdisciplinary doctorates) - Joint applications for research funding with colleagues in other fields   INTERNATIONAL COLLABORATION   - Leading role in international scientific organisations, such as member of the organising committee of an internationally-renowned conference - Management of an international research project (e.g. VLIR-UOS, EU,…) or research applications as a member of international consortia - Research funding obtained from international sources - New bilateral collaboration projects - Development, continuation and reinforcement of an international network for high-quality collaboration and exchange - Publications in the context of international collaborations - Joint doctorates - Supervision of international exchange doctoral students - Patents with international co-applicants   ACADEMIC COMMITMENT   - Participation in research-related policy committees - Leading role as an evaluator or expert in the researcher’s field (e.g. panel chair, member of a “high level group”) - (Co-)authorship of reports or opinions on research or innovation policy - Leading or strategic role based on the researcher’s own general research expertise, for example in Data Protection Authority, the Belgian Federal Agency for Medicines and Health Products, an international scientific association - Membership in external assessment committees   SCIENTIFIC IMPACT   - Keynote speaker at the most renowned conferences in the field - Edition/circulation of a book publication by a renowned academic publisher - Editor (by invitation) of a special issue of a journal or scientific book series - Member in or chair of assessment or evaluation committees outside of Ghent University - Scientific recognition or award for making a paradigm shift in the field - Number of downloads of (open access) papers - Number of users/downloads of (open) datasets - Normalised citation impact of SSCI and SCIE publications during the reference period - Highly cited papers - Recognition as a Highly Cited Researcher   SOCIETAL or ECONOMIC IMPACT   - Development of a valorisation strategy for research results - Preparation/creation of spin-offs - Creation of an endowed chair - Being part of an IOF (Industrial Research Fund) business development centre as a steering committee member with a contribution to valorisation work - Research projects in collaboration with non-academic partners (industry, government, private non-profit, etc.), strategic basic research projects, policy-related research projects, etc. - Setting up interdisciplinary innovation platforms or clusters that jointly federate different research groups - International public profile in terms of influencing professional practice and policy - Circulation/distribution of a book publication with a general publisher (other than compulsory course material) - Research projects in collaboration with partners in developing countries - Development of a training package or awareness campaign (within or outside Ghent University, or for example in the context of research collaborations and/or development projects with academic partners in the South) - Structural cooperation with Ghent University public platforms (GUM, Krook, etc.) - Structural involvement of research stakeholders (living labs, co-creation processes, citizen science, etc.) - Strategic approach for communication via public media (e.g. opinion pieces, blog posts, contributions to Wikipedia, ikhebeenvraag.be, etc.) - Participation in Open Innovation projects - Patent applications submitted and patents granted - Clinical studies - Participation (organisation, keynotes, etc.) in relevant public events, stakeholder events, Science Communication, Science Shop activities, etc. - Manuals for schools and other education providers - Organisation of an exhibition/show - Number of policy reports and opinions - Altmetrics | From Vision Statement:  Strike the right balance between indicator-driven (quant) and peer-review-driven (qual) assessment methods:   1. Choose the method or blend of methods that matches assessment objective 2. Consider the intended impact (academic, economic, societal) 3. Account for diversity between disciplines 4. Evaluating the quality of research cannot be based on one indicator, but reduce the administrative burden on researches by only requesting information that will actually be used in assessment 5. Evaluation criteria are shared to all stakeholders in advance 6. There are sufficient experts on the evaluation committee to adequately assess the quality of the research |
| Federation of Finnish Learned Societies, Finland  Good Practice in Researcher Evaluation. Recommendations for the responsible evaluation of a researcher in Finland [51]  February 2020 (30 pages) | Academic | Generated by 50 research-related organizations plus two rounds of feedback, first the entire Finnish research community, then public consultation | Research funding, supervision and leadership   - leadership in the research community (e.g. department head or research team leader) - duties in the administration or working groups of universities and research organisations - significant research funding (applications made and funding received) - mentoring of post-doc researchers - acting as an officially designated instructor for undergraduate and postgraduate students   Awards and honours   - awards and honours for scientific, artistic or research merits or on the basis of academic career   Scientific merit   - referee duties for scientific journals - acting as a pre-examiner or opponent of a dissertation; membership of doctoral dissertation boards (abroad) - ievaluation of scientific / artistic competence (e.g. title of docent) - participation in the national or international peer review of funding applications as an expert   Networking and community development   - Memberships and elected positions in scientific communities such as learned societies and academies - membership in a national or international expert, evaluation or steering group and other expert duties - participation in scientific editorial work (e.g. membership of scientific publication series’ and journals’ editorial boards and editorial positions, including editor-in-chief) - major international invitation lectures - willingness and ability to work in diverse groups and work communities - activities and positions promoting and encouraging research cooperation   Scientific and societal impact   - merits in producing and sharing research and information materials - merits in utilising research results - invention announcements, patents, and other merits in promoting commercialisation (e.g. spin-offs and trademarks) - merits in science communication and performing as an expert in the media - principal public positions of trust, expert positions and assignments (including research-based policy-advice duties) - duties as an invited scientific expert - collaborative projects with major research actors including applied research projects | General Principles  Transparency, integrity, fairness, competence, diversity  Recommendations   - Quantitative indicators can be used to support qualitative peer review of scientific activity. Peer review should be the primary approach for evaluating individual researchers. - Publication metrics should be based on data that is relevant for the unit of assessment. The known limitations of the data should always be disclosed. - Being as open and transparent as possible in data collection, analytical processes and results is necessary. Those being evaluated should, as far as possible, be able to check both the data used and the results of the analysis. - Disciplinary differences and interdisciplinarity should be taken into account in the application of publication metrics. - The indicators used in assessment should be chosen to support the aims of the evaluation. - Results should be reported with an accuracy relevant for the unit of assessment, methods and the data. Inapplicable indicators should not be reported. - Specific expertise is needed in the production and interpretation of publication metrics. - Organisations committed to this recommendation should provide sufficient resources and expertise needed for producing and interpreting publication metrics. - Organisations should offer training for responsible use of publication metrics for their faculty and staff. - Organisations committed to this recommendation should name the responsible party in their organisation who can be contacted in cases of irresponsible use of publication metrics.   Consider characteristics that differ by discipline:   - differences in publishing practices in terms of metrics, different evaluation models and transparency; - different forms of research output (e.g. research methods, software and artistic output); - the prevailing size of research teams in the research field and the division of labour among researchers and their interdependence in research work; - principles defining authorship; - international penetration of the research field and opportunities for international research cooperation; - different evaluation practices within interdisciplinary and multidisciplinary projects and the tensions between them; - forms of societal interaction; - opportunities and requirements for commercial cooperation; and - the role of co-development and civic science in research. |
| Universities Norway Consortium, Norway  Evaluation of research careers fully acknowledging Open Science practices: rewards, incentives and/or recognition for researchers practicing Open Science [52]  July 2017 (32 pages) | Academic | Recommendations to promote the practice of Open Science | RESEARCH OUTPUT  Research activity  Pushing forward the boundaries of open science as a research topic  Publications  Publishing in open access journals; Self-archiving in open access repositories  Datasets/Outputs  Using the FAIR data principles; Adopting quality standards in open data management and open datasets; Making use of open data from other researchers  Open source  Using open source software and other open tools; Developing new software and tools that are open to other users  Funding  Securing funding for open science activities  RESEARCH PROCESS  Stakeholder engagement/citizen science  Actively engaging society and research users in the research process; Sharing provisional research results with stakeholders through open platforms (e.g. Arxiv, Figshare); Involving stakeholders in peer review processes  Collaboration/Interdisciplinarity  Widening participation in research through open collaborative projects; Engaging in team science through diverse cross-disciplinary teams  Research integrity  Being aware of the ethical and legal issues relating to data sharing, confidentiality, attribution and environmental impact of open science activities; Fully recognizing the contribution of others in research projects, including collaborators, co-authors, citizens, open data providers  Risk management  Taking account of the risks involved in open science  SERVICE/LEADERSHIP  Leadership  Developing a vision and strategy on how to integrate OS practices in the normal practice of doing research; Driving policy and practice in open science; Being a role model in practicing open science  Academic standing  Developing an international or national profile for open science activities; Contributing as editor or advisor for open science journals or bodies  Peer review  Contributing to open peer review processes; Examining or assessing open research  Networking  Participating in national and international networks relating to open science  RESEARCH IMPACT  Communication and dissemination  Participating in public engagement activities; Sharing research results through non-academic dissemination channels; Translating research into a language suitable for public understanding  IP (patents, licenses)  Being knowledgeable on the legal and ethical issues relating to IPR; Transferring IP to the wider economy  Societal impact  Evidence of use of research by societal groups; Recognition from societal groups or for societal activities  Knowledge exchange  Engaging in open innovation with partners beyond academia | Recommendations:   1. To change the culture and further engage the entire researcher community in the practice of Open Science a more comprehensive recognition and reward system incorporating Open Science must become part of the recruitment criteria, career progression and grant assessment procedures for researchers at all levels 2. Where needed, there should be a review of ERA policies, ERA roadmaps and National Action Plans through the lens of Open Science. If necessary, policies must be updated in order to ensure compatibility with Open Science. 3. Encourage and incentivise researcher participation in Open Science 4. The assessment of researchers during recruitment, career progression and grant evaluation should be structured to encompass the full range of their achievements including Open Science. |
| University College London  Academic Careers Framework [53]  July 2018 (28 pages) | Academic | To help plan and support career development  and recognise achievements | - References from group leader, supervisors and immediate collaborators - Refereed conference posters/papers - Peer-reviewed journal articles, book chapters - Peer-reviewed cultural, artistic or design outputs, as appropriate to the discipline - Occasional reviewer for research-focused journals - Descriptions of impact activities - Participation in policy-focused meetings or events; engagement with UCL Public Policy activities e.g. undertaking a policy placement; evidence of scoping and responding to policy stakeholder needs - Effective supervision and mentoring of PhD students - Proactive engagement with research development issues across the faculty - Supervisor or second supervisor experience of research students - Findings supported/invitations extended to disseminate these at conferences and similar - Academic references from across discipline community - Paper co-authored with collaborator with evidence of impact within the discipline - Significant cultural, artistic or design outputs, as appropriate to the discipline - Conference speaker invitations, including as a consequence of submitting proposals to conference panels - Regular reviewer for research-focused journals - Collaborator in research grant application - Successfully co-organised event aimed at an external audience. - Personal contribution to initiative to contribute to equalities and diversity objectives within field - Contributions to Open Source software, large scale computing projects - Successful supervision to completion of doctoral students - Evidence of positive impact of mentoring of colleagues, including that they have met their career development goals - Research fellowship or award from UK or international funding body - Sustainable research project with funding successes in a competitive context and at a level appropriate to the discipline - Whole monographs, including as editor, where appropriate to the discipline - Cultural, artistic or design outputs with funding successes or other appropriate evidence of impact, in a competitive context - References from national and/or international subject community, including leading figures - Editorial board member of a significant journal in your discipline - Significant income generated through external clinical trials - High quality research outputs from collaborative research projects of significant standing. - Invited speaker, including keynote, at significant national and/or international events, or invitations to write review articles - Personal impact within cross-disciplinary programmes - Proactive engagement with national policy working groups - Peer reviewer for grant schemes both nationally and internationally - Lead role in creation of a new research facility or group (where appropriate to the discipline) - Sustained contribution to initiative to contribute to equalities and diversity objectives within field, with evidence of impact - Evidence of impact of research on clinical practice or pathways (translation) - Evidence of impact of research on clinical guidelines (e.g. NICE, Cochrane) - Mentoring of research supervisors with evidence of impact - Sustained track-record of income generation to support own group or field of work - Chair of departmental or faculty research committee - Outstanding sustained quality of research outputs, including those based on collaborations where appropriate - Leadership of a major research field or group with track record of securing competitive grants, including leadership within a wider team where this is appropriate to the discipline context - Editor of a significant research journal or book series - Regular keynote speaker invitations at conferences attracting international participation - Introduction of new research methods or approaches to the discipline - Leading role in significant networks or associations relevant to discipline (including e.g. Learned Councils) - Regular invitations to deliver research or analysis by external organisation(s) and/or member of policy groups of significant standing - Success in a major multi-group collaborative funding opportunities - Internationally significant policy positions on standards within discipline and similar. - Leadership of successful initiative to contribute to equalities and diversity objectives within field - Evidence of substantial impact of research on clinical practice or pathways (translation) - Evidence of substantial impact of research on clinical guidelines (e.g. NICE, Cochrane) - Sustained recognition by policy professionals and stakeholders | - Publish research findings and proposals - Disseminate research findings at conferences and similar - Participate in research-related enabling activities, for instance adding value to a cross-disciplinary network, journal reviewing - National and/or international engagement - Engagement with UCL Public Policy strategic activities e.g. policy round tables, the production of policy-focused outputs; evidence of building networks or partnership with policy stakeholders; testimonials from policy professionals; adopting co-production methods - Provide peer review, e.g. by serving on peer review committees - Work towards an independent research reputation (or the research reputation of a collaborative team) and recognition of achievement nationally/internationally - Significant contribution to external engagement with a view to enhancing research impact - Enabling scientific input (and output) on research platforms |
| University Medical Centre Utrecht, Netherlands  Guide for reviewers/evaluators [54]  No date (6 pages) | Academic | Framework of indicators for research assessment | Collaboration with stakeholders   - Description of stakeholders - Overview of meetings held with stakeholders - Demonstrable interest of stakeholders: staff exchange; shared publications; public-private partnerships; shared IP; memberships of advisory councils or other manifestations of collaborations with public parties   Setting research priorities   - Provide the mission, a text that answers the question why the unit does what it does - Description of the questions that are being pursued and how the answers will help us further. The questions might for example relate to an “unmet medical need”; relate to a biomedical or healthcare problem; or might involve a new and promising technology or research method; etc   Posing the right questions   - Demonstrate how the main research question fits in with existing knowledge, for example by referring to (systematic) reviews; to (multidisciplinary) roadmaps or to research agendas - Describe which stakeholders were involved, and how, in formulating research questions   Incorporation of next steps   - Possible users of research findings are demonstrably involved in the project, e.g. other (clinical) research groups, general practitioners, nurses, small and medium enterprises, pharmaceutical and medtech companies, etc - Presence of a dedicated ‘business developer’ or other demonstrable support for innovation and valorization - Funding from companies, charities, patient organizations, health insurers; etc - Membership of (guideline) committees, policy panels; lectures for policy makers and other stakeholders; publications in “grey literature”; coverage in general media; etc   Design, conduct, analysis   - Description of statistical and methodological support - Number of DEC and METC applications; - If available: include results from JCI research tracers   Regulation and management   - Availability of data management plans - Publication of raw data; or the availability of data for external use - Pre-registration of protocols (both in pre-clinical and clinical research); - Reproduced publications and/or reproduction efforts; - Clinical trial registration and publication   Research products for peers  Describe the three most important research products for peers, consisting of key publications other forms of research output, such as scientific/scholarly books, instruments, infrastructure, intellectual property, datasets, software tools or designs that the unit has developed; number of dissertations  Research products for societal target groups  Provide the three most important examples of research products for societal target groups, e.g. reports (for example for policymaking); articles in professional journals for non-academic readers; other outputs (instruments, infrastructure, intellectual property, datasets, software tools or designs that the unit has developed) for societal target groups; or outreach activities, for example lectures for general audiences and exhibitions.  Use of research products by peers  Provide the three most important examples on how research products are being used, e.g. in terms of citations for selected articles; the use of datasets, software tools, etc. by peers; use of research facilities by peers  Use of research products by societal groups  Provide the three most important examples of use of research products by societal groups, e.g. implementation of new treatments/acceptance as standard of care (also by health insurers); incorporation of products in guidelines; use of research facilities by societal parties; projects in cooperation with societal parties; contract research  Marks of recognition from peers  Provide the three most important examples of recognition from peers, e.g. science awards/scholarly prizes; research grants awarded to individuals; invited lectures; membership of scientific committees, editorial boards, etc.  Marks of recognition from societal groups  Provide the three most important examples of marks of recognition from societal groups, e.g. public prizes, appointments/positions paid for by societal parties, membership of civil society advisory bodies; valorisation funding |  |
| University of Bath, United Kingdom  Principles of research assessment and management [55]  [https://www.bath.ac.uk/ corporate-information/](https://www.bath.ac.uk/ corporate-information/ principles-of-research- assessment-and-management/)  [principles-of-research-](https://www.bath.ac.uk/ corporate-information/ principles-of-research- assessment-and-management/)  [assessment-and-management/](https://www.bath.ac.uk/ corporate-information/ principles-of-research- assessment-and-management/)  Accessed March 2021 | Academic | Principles that outline approach to research assessment and management, including the responsible use of quantitative indicators | --- | Centred on expert judgment  Quantitative indicators can be used to inform judgements and challenge preconceptions, but not to replace expert judgement.  Set in broader environment  Be aware of the potential for research assessment and associated metrics to reflect or introduce bias  Supported by reliable data  The aim is to avoid placing undue significance on quantitative differences out of context. Research quality is multifaceted and cannot be captured by a single indicator used in isolation.  Tailored: one size does not fit all  Disciplinary differences in research inputs, processes and outputs have to be taken into account. Evaluation should also be tailored to the scale of the research activity being assessed.  Transparent  Assessment criteria and any quantitative data used must be transparent and available (on request) to those being assessed. Those conducting assessments must disclose the data sources used and ensure that researchers can review and correct data about their work. |
| University of California, Irvine, United States  Identifying Faculty Contributions to Collaborative Scholarship [56]  [https://ap.uci.edu/faculty/ guidance/collablist/](https://ap.uci.edu/faculty/guidance/collablist/) | Academic | Framework by which to assess research contributions | Conceptual Contributions   - Contribute the key idea behind the work - Have critical insight that breaks a conceptual logjam - Create theoretical ideas or frameworks - Contribute relevant literature   Methodological Contributions   - Bring expertise in a particular research approach - Develop or share relevant software for modeling or analysis - Bring statistical expertise - Create visualizations that help create understanding during analysis - Provide data curation   Resource Contributions   - Help obtain grant funding - Contribute funds from an existing source - Possess relevant specialized skills (either self or staff) - Build or provide access to specialized equipment or facilities - Provide critical materials (e.g., cell lines) - Provide existing data sets - Recruit research participants - Especially if special populations are required - Establishing relations to organizations that link to relevant populations   Project Level Contributions   - Provide overall project administration, leadership - Especially important for geographically distributed projects - Especially important for cross-disciplinary collaborations - Be a liaison to a key community or organization - Introduce or refer important people to team members - Support team building, getting researchers to speak the same language, trust each other, mentor   Dissemination Contributions   - Take leadership in creating the papers - Do significant work in editing papers for clarification and transparency - Create and give presentations - Translate the research to practitioners and the public - Create useful visualizations of data or the models for others to understand - Commercialize the technologies, acquire patents |  |
| American Society of Microbiology  Rescuing Biomedical Research [57]  <https://rescuingbiomedicalresearch.org/>  PNAS article April 2014  Alberts et al. 2014;111:5773-5777  Rescuing US biomedical research from its systemic flaws  <http://rescuingbiomedicalresearch.org/> wp-content/uploads/2015/07/PNAS-2014-Alberts-5773-7.pdf | Professional society | Platform to discuss solutions to problems such as those addressed in the PNAS article entitled |  | PNAS recommendations for improving evaluation criteria:  The tools used to judge past performance should be sharpened to identify the strongest candidates for support. The qualitative aspects of each candidate’s major scientific achievements should receive more emphasis than the numbers and venues of publications. Evaluation criteria should also put a higher priority on the quality, novelty, and long-term objectives of the project than on technical details.  Review guidelines should be appropriately  adjusted for young scientists to promote  the funding of thoughtful proposals that  reveal ingenuity and promise findings with  potentially broad implications. The criteria  used to evaluate the NIH Director’s New  Innovator Award set useful standards. |
| Austrian Science Fund  Application Guidelines for Stand-Alone Projects [58]  November 2020 (23 pages) | Funder | Recognizes a diverse range of research outcomes and requires grant applicants to provide information on various research achievements | Required description of previous research achievements:  Academic publications: list of no more than ten of the most important published or accepted academic publications (journal articles, monographs, edited volumes, contributions to edited volumes, proceedings, etc.); for each publication, if available, either a DOI address or another persistent identifier must be indicated. In accordance  Application Guidelines Stand-Alone Projects November 2020 11 with the San Francisco Declaration on Research Assessment (DORA), journal-based metrics like the journal impact factor should not be included.  Additional research achievements: list of no more than ten of the most important research achievements apart from academic publications (such as awards, conference papers, keynote speeches, important research projects, research data, software, codes, preprints, exhibitions, knowledge transfers, science communication, licenses, or patents). | --- |
| Australian National Health Medical Research Council  Guide to NHMRC Peer Review [59]  2018 (19 pages) | Funder | Requires consideration of a broad range of measures that affect the assessment of an applicant’s research achievement | NHMRC requires consideration of a broad range of measures that affect the assessment of an applicant’s research achievement. These include both quantitative and qualitative measures, such as  the scientific value of publications and influence on current dogma, policy or practice.  The value of the research achievement is indicated by:   - the number of citations of individual publications - success in obtaining peer reviewed grants - contribution to translational outcomes such as patents - commercial output - public policy or implementation of change in practice - invitations to conferences - mentoring, leadership - speaking engagements - numbers and types of prizes and awards - contribution to the research community. | Section 4.8  Use of Impact Factors and other metrics  Peer reviewers should take into account their expert knowledge of their field of research, as well as the citation and publication practices of that field, when assessing the publication component of an applicant’s track record. Track record assessment should take into account the overall impact, quality and contribution to the field of all of the published journal articles from the grant applicant, not just the standing of the journal in which those articles are published.  It is not appropriate to use publication metrics such as Journal Impact Factors. NHMRC is a signatory of DoRA and adheres to the following recommendations, as outlined in DoRA, for its peer review processes:  Do not use journal-based metrics, such as Journal Impact Factors, as a surrogate measure of the quality of individual research articles, to assess an individual scientist’s contributions, or in hiring, promotion, or funding decisions.  Be explicit about the criteria used in evaluating the scientific productivity of grant applicants and  clearly highlight, especially for early-stage investigators, that the scientific content of a paper is much more important than publication metrics or the identity of the journal in which it is published.  For the purposes of research assessment, consider the value and impact of all research outputs (including datasets and software) in addition to research publications, and consider a broad range of impact measures including qualitative indicators of research impact, such as influence on policy and practice.  Citation metrics such as h-index, m-index or g-index used in isolation can potentially  be misleading when applied to the peer review of publication output, as they do not describe the impact, importance or quality of the publication(s). They are also dependent on the citation practices of different research fields and are therefore not a reliable comparative measure across research fields. Such metrics must be considered within a broad range of measures as outlined above and used with caution. |
| Netherlands research funders (VSNU, NFU, KNAW, NWO, and ZonMw)  Strategy Evaluation Protocol 2021-2027  March 2020 [60] (48 pages)  ALSO  Room for everyone's talent towards a new balance in the recognition and rewards of academics (7 pages)  Infographic aimed at researchers to create awareness of the Strategy Evaluation Protocol and explain that a change in assessment is needed to focus on research quality rather than metrics | Funder | Criteria and process for evaluating research units every six years (institutes, departments, research groups) | Appendix E. Merit and metrics  The research unit should take into account that it is not allowed to use the Journal Impact Factor in a SEP evaluation. The Journal Impact Factor was not created as a measure of the scientific quality of research in an article. It has a number a number of well-documented deficiencies as a tool for research assessment11. The use of the h-index is advised against because 1) it is sensitive to age and experience (so young scholars always have low h-index values), 2) it is not field-normalised, which makes comparison across fields – sometimes even within fields – based on the h-index impossible and 3) it is an author-level metric, while SEP assessments evaluate research units.  For each of research quality and societal relevance, there must be quantitative and/or qualitative evidence of:  Demonstrable products  Demonstrable use of products  Demonstrable marks of recognition  Options for Research Products   - Open access journal articles and reviews - Open access books, monographs or chapters - Editorship of volumes or special issues - Digital infrastructure and databases - Presentations and conference proceedings - Designs (tangible outputs or simulations) - Data sets and software - Guidelines - Patents or licenses - Films, documentaries, exhibitions - Media or communication for lay persons (e.g. books, infographics, digital applications, lectures, blogs) - Other: research materials, instruments, infrastructure, websites, lectures delivered at research conferences, organisation of scientific conferences, films, commissioned reports, annotations.   Options for Use of Products   - Reviews - Use of data sets, software, or scholarly literature - Citations of articles, books or other products - Projects in collaboration with societal parties - Contract research - References in professional and public domains   Options for Recognition   - Research grants awarded - Grants awarded to major collaborative initiatives - Prestigious research prizes or honours - Financial and material support by society - Appointments or membership in prestigious scientific councils or committees | Three assessment criteria  Research quality:  the quality of the unit’s research over the past six-year period is assessed in its international, national or – where appropriate – regional context. The assessment committee does so by assessing a research unit in light of its own aims and strategy. Central in this assessment are the contributions to the body of scientific knowledge. The assessment committee reflects on the quality and scientific relevance of the research. Moreover, the academic reputation and leadership within the field is assessed. The committee’s assessment is grounded in a narrative argument and supported by evidence of the scientific achievements of the unit in the context of the national or international research field, as appropriate to the specific claims made in the narrative. The protocol explicitly follows the guidelines of the San Francisco Declaration on Research Assessment  Societal relevance:  the societal relevance of the unit’s research in terms of impact, public engagement and uptake of the unit’s research is assessed in economic, social, cultural, educational or any other terms that may be relevant. Societal impact may often take longer to become apparent. Societal impact that became evident in the past six years may therefore well be due to research done by the unit long before. The assessment committee reflects on societal relevance by assessing a research unit’s accomplishments in light of its own aims and strategy.  Viability:  the extent to which the research unit’s goals for the coming six-year period remain scientifically and societally relevant is assessed. It is also assessed whether its aims and strategy as well as the foresight of its leadership and its overall management are optimal to attain these goals. Finally, it is assessed whether the plans and resources are adequate to implement this strategy. The assessment committee also reflects on the viability of the research unit in relation to the expected developments in the field and societal developments as well as on the wider institutional context of the research unit. |
| European Commission  Evaluation of Research Careers Fully Acknowledging Open Science Practices: Rewards, incentives and/or recognition for researchers practicing Open Science [61]  July 2017 (80 pages) | Funder | Recommendations on how to incent and reward researchers for Open Science | Repeat of the following: SEE:  Universities Norway Consortium, Norway  Evaluation of research careers fully acknowledging Open Science practices: rewards, incentives and/or recognition for researchers practicing Open Science  July 2017 (32 pages) | Open Science offers researchers the means for greater transparency, reproducibility, dissemination and transfer of new knowledge. OS provides greater access to data and publications which can improve the effectiveness and increased productivity of researchers (allowing more research from the same data). In an open environment there can be a more accurate verification of research results. These are examples of good reasons for researchers to practise OS. |
| Wellcome Trust  Open Access 2020 Policy [62]  November 2018 (2 pages) | Funder | To promote Open Science | We expect our researchers to publish their findings freely online as high-quality, peer-reviewed research articles, monographs and book chapters  All research articles supported in whole or in part by Wellcome must be:  - made freely available through PubMed Central (PMC) and Europe PMC by the official final publication date, and  - published under a Creative Commons attribution licence (CC-BY).  - post preprints of their completed manuscripts  All research articles supported in whole or in part by Wellcome must include a statement explaining how other researchers can access any data, original software or materials underpinning the research | Providing free, online access to published research will maximise the availability and usability of publications, and make sure the research we fund can be built upon  We will no longer cover the costs of OA publishing in subscription journals. Grant applicants cannot ask for these costs in their grant application, and grantholders will not be allowed to use their grant funds to pay for these costs. |
