## Supplemental Appendix 5 for "DORA-compliant measures to assess research quality and impact in biomedical institutions: review of published research, international best practice and Delphi survey"

**APPENDIX 5. Compiled research assessment measures**

**1.0 Relevance of Research Program**

NOTE: One or more of the following may be relevant dependent on research discipline

| 1.1 | Research addresses societal needs (e.g. reveals or responds to unmet needs) |
| --- | --- |
| 1.2 | Research advances existing applied and/or theoretical knowledge |
| 1.3 | Research demonstrates a logical trajectory of research (i.e. projects successively build on completed research) |
| 1.4 | Research plan is innovative |
| 1.5 | Research plan is feasible |
| 1.6 | Synergy of research plan with organizational priorities (e.g. UHN, Research Institutes, Programs) |

**2.0 Challenges to Productivity of Research Program**

NOTE: Factors to consider when assessing candidates so that evaluations/comparisons are based on equitable support; does NOT include interruptions due to maternity leaves, caregiver responsibilities, health leaves, etc.

| 2.1 | Qualitative description by researcher of challenges faced and mitigating strategies applied |
| --- | --- |
| 2.2 | Infrastructure provided by organization or sponsors (e.g. space) |
| 2.3 | Dedicated resources provided by organization or sponsors (e.g. lab manager, administrative support) |

**3.0 Team/Open Science**

NOTE: Collaboration, sharing of research for all to access

| 3.1 | Evidence of local, regional, national or international team science (e.g. formation of clinical network or inter-/multi-disciplinary research group) |
| --- | --- |
| 3.2 | Participation as co-investigator or mentor on other research teams |
| 3.3 | Co-production of research with those outside academia (e.g. policy-makers, patients/family, public) |
| 3.4 | Registration of planned research (e.g. trials, syntheses) |
| 3.5 | Open publication/sharing of data sets, software, code, biological materials (e.g. plasmids, cell lines, model organisms), other materials, tools, reports or publications |

**4.0 Funding**

NOTE: Acquisition of resources to conduct research

| 4.1 | Evidence of research independence (e.g. funding as PI) or fundamental role (e.g. biostatistician, qualitative researcher) in peer-reviewed research funding |
| --- | --- |
| 4.2 | Proportion of peer-reviewed research funding that is from Tri-Council sources |
| 4.3 | Attempts to capture peer-reviewed research funding (number of submitted applications) |
| 4.4 | Attempts to capture non-peer-reviewed research funding (e.g. number of submitted applications, networking activities) |
| 4.5 | Peer-reviewed, competitive salary support (e.g. New Investigator, Chairs) |
| 4.6 | Number of peer-reviewed applications funded |
| 4.7 | Number of peer-reviewed applications funded but declined due to overlap in scope |
| 4.8 | Number of peer-reviewed non-competitive, non-peer-reviewed and/or industry-sponsored grants, research agreements, clinical trials |

**5.0 Innovations**

NOTE: Outputs of research NOT including publications, which are considered separately; may be specific to discipline

| 5.1 | Commercialization of technology (e.g. software, drugs, devices), launch of companies, invention disclosures, patent applications, issued patents, licenses, etc. |
| --- | --- |
| 5.2 | Creation of cohorts or registries |
| 5.3 | Clinical tests, algorithms or statistical models registered or available in a repository |
| 5.4 | Validated questionnaires or instruments |
| 5.5 | Development of, contribution to, or use of one’s research in policies, guidelines, standards or programs |
| 5.6 | Development and sharing of a novel theory, model or framework |
| 5.7 | Novel research approaches (e.g. development of new research methods) |
| 5.8 | Other forms of creative achievement or outputs relevant to discipline (e.g. physical simulation, performance art) |

**6.0 Dissemination – Publications**

NOTE: counts should not be used alone; scientific contributions and impact should receive more emphasis than the numbers and venues of publications

| 6.1 | Quality of the content of publications (qualitatively assessed based on description by researcher of contribution) |
| --- | --- |
| 6.2 | Number of peer-reviewed journal publications, principal/co-principal author(s) [**when applying this indicator, tailor to discipline: or other positions reflecting proportion of contribution; for example, first three authors] |
| 6.3 | Number of peer-reviewed journal publications, senior responsible author(s) [**when applying this indicator, tailor to discipline; for example, senior author, corresponding author] |
| 6.4 | Number of peer-reviewed journal articles published via open access (i.e. freely available to all, required by Tri-Council funding agencies) |
| 6.5 | Number of books, principal author or editor |
| 6.6 | Number of book chapters |
| 6.7 | Number of preprints |
| 6.8 | Other types of peer-reviewed and non-peer reviewed publications (e.g. editorials, commentaries, guidelines, task force reports for government or public organizations) |

**7.0 Dissemination - Presentations, Media, Social Media and Creative Approaches**

| 7.1 | Invited presentations at academic or professional society meetings |
| --- | --- |
| 7.2 | Accepted refereed meeting proceedings, abstracts, papers, posters or presentations |
| 7.3 | Presentations or meetings with healthcare professionals, policy-makers, community groups or general public |
| 7.4 | Media mentions (e.g. text or broadcast via television, radio, newspaper, magazine) |
| 7.5 | Number of social media posts/appearances (e.g. tweets, blogs, podcasts, videos), or social media roles (e.g. curator, editor) or reach (e.g. number of followers, likes, downloads, page views) |
| 7.6 | Creative approaches to knowledge mobilization (e.g. research-based drama/film, animation, art installations) |

**8.0 Evidence of Impact**

NOTE: impact on science, practice, policy, health systems, society; NOT publications, which are considered separately

| 8.1 | Researcher reputation (e.g. leadership role in local/provincial/national/international committees, organizations, or conference steering groups) |
| --- | --- |
| 8.2 | Distinctions, credentials, honours and awards, funded and unfunded (e.g. vetted fellowships or membership in scholarly societies such as Canadian Association of Health Sciences) |
| 8.3 | Leadership or co-leadership of clinical translation through Phase 1 to 3 clinical trials |
| 8.4 | Contributions to organizational, government or health system policy development or changes (e.g. based on professional expertise in an advisory capacity or influence of research on policy-makers/policy) |
| 8.5 | Demonstrated use of research outputs by other researchers, clinicians, organizations, government, patients/families |
| 8.6 | Social, cultural, population, environmental or economic returns from use of research outputs/products and associated outcomes (e.g. identification of health inequities among social or cultural groups, translation of research into commercial products, improved clinical outcomes or quality of life among populations) |
