## Supplemental Appendix 6 for "DORA-compliant measures to assess research quality and impact in biomedical institutions: review of published research, international best practice and Delphi survey"

**APPENDIX 6. Research assessment guidance: principles, responsibilities and processes**

**PRINCIPLES**

- Create a more porous research culture to promote interdisciplinary approaches, and enable more mobile and flexible research careers
- Create a more flexible template for the academic CV so that researchers can best highlight their unique contributions
- Scrutinize indicators regularly and update them (e.g. every 6 years so that indicators are updated at least once during the review period of a Scientist); refine research assessment processes through iterative feedback
- Avoid misplaced concreteness and false precision; instead, use multiple indicators to provide a more robust and pluralistic picture; one size is unlikely to fit all – a mature research system needs a variable geometry of expert judgement, quantitative and qualitative indicators
- Quantitative evaluation (metrics) should support qualitative, expert assessment; a simple counting of publications or grants/grant value without assessment of their quality is not sufficient and may be misleading
- Have individuals highlight and articulate their most meaningful contributions by describing the quality, significance and impact of their scholarship and specify the level of impact: individual, community, system, population to help reviewers assess its merits
- Base assessment of individual researchers on a qualitative judgement of their portfolio
- Keep research productivity assessment data collection and analytical processes equitable, open, transparent and simple
- Account for variation by field in funding (balance of industry-sponsored versus competitive, peer-reviewed sources) publishing practices, evaluation models, size of research teams and division of labour, principles defining authorship, etc.
- Take a big picture or portfolio view toward researcher contributions
- Productivity and quality should rise as faculty move through the academic ranks; members with a lower percentage research (10%) and rank (Assistant Professor/Affiliate Scientist) are expected to have at least one product or process of scholarship or item of dissemination per reporting year; members with a higher percentage research and/or higher academic rank (Associate or Full Professor/Senior Scientist) are expected to have more items and/or of increasing quality (broader scope, greater impact) of scholarship [**this may vary for some disciplines]
- All scholarly endeavors are time consuming activities, and during development, productivity/outputs may appear low; thus, individuals must describe the process of scholarship and demonstrate appropriate progress

**RESPONSIBILITIES**

- Institutional leaders should develop a clear statement of principles on their approach to research management and assessment (that may be discipline-specific or tailored to discipline)
- Institutions committed to these principles should name the responsible party in their organisation who can be contacted in cases of irresponsible use of publication metrics
- Research managers and administrators should champion these principles and the use of responsible evaluation within their institutions
- HR managers and recruitment or promotion panels should be explicit about the criteria used for appointment, annual or periodic performance review, and promotion decisions
- Individual researchers should be mindful of the limitations of particular indicators in the way they present their own CVs and evaluate the work of colleagues
- Individual researchers are responsible for providing narrative descriptions of the quality, context, and impact of their research, individual research products, and other aspects of research without relying on surrogate metrics. The person best-placed to articulate the importance of the research is the researcher themselves.

**REVIEW COMMITTEES**

**Hiring, compensation, re-appointment, promotion, consideration for leadership roles, etc.**

- Review or selection committees must include disciplinary peers (research leaders and researchers) representing each department or discipline, elected for two-year terms
- Members may serve multiple terms as regular members; however, no member shall serve two consecutive terms in this capacity
- Assemble diverse committees reflecting inclusivity, diversity, equity and ability—across gender, seniority, cultures, and under-represented minority populations—to bring a range of perspectives and experiences into decisions
- Thoughtful expert evaluation is an institutional priority and requires time. To allow this, those serving on review committees should either have reduced duties elsewhere, or should not be expected to have the same level of research output as others achieving the same assessment. This should also be applied to other committee service with heavy review requirements outside committee meetings (for example, research ethics board or animal use committees).

**REVIEW PROCESSES**

- To help members plan research trajectories and prepare for review, the Research Institute or Unit should provide a manual that describes the process, and provides examples of lower and higher quality research activities for each discipline
- To facilitate success, each Research Institute or Unit Chair shall:
  - Review with each member their responsibilities and expectations
  - Meet with each member annually to discuss their annual report; and performance quality, progress and trajectory
  - Discuss career goals, and offer mentorship and other supports
  - Discuss merit recommendations
  - Jointly agree on an action plan to address deficiencies
- Allow those evaluated to verify data and analysis
- Assessment categories:
  - Excellent: performance that is: a) functioning beyond commendable for their rank and/or percentage of position description, and/or b) distinguishing and expanding skills/learning opportunities/stewarding our people, our work and service
  - Commendable: performance that is beyond Acceptable will be distinguished for merit
  - Acceptable: performance demonstrates a significant deficiency in one domain of evaluation, but performs well in the other domains, or when the faculty member overall performs below expected for rank, but remains within the acceptable range
  - Unacceptable: Academic performance is unsatisfactory and unacceptable

**PROMOTING DORA PRINCIPLES**

- Create an action plan to implement, promote and routinely review these principles
- Promote DORA principles organization-wide (e.g. employ infographics on DORA web site)
  - Rethinking Research Assessment: Ideas for Action
  - Rethinking Research Assessment: Unintended cognitive and systems biases
- Provide education/training to research leaders and researchers about these principles, and how to assess (leaders) and describe (researchers) research productivity/contributions
- Incentivize and reward a broader range of academic activities
