## Supplemental Appendix 7 for "DORA-compliant measures to assess research quality and impact in biomedical institutions: review of published research, international best practice and Delphi survey"

**APPENDIX 7. Respondent ratings of all measures**

| Measures | Rated important (%) | Consensus to retain |
| --- | --- | --- |
| Relevance of Research Program |  |  |
| Research directly addresses or has the potential to improve the health and healthcare of Canadians | 54.2 | ✓ |
| Research advances existing applied and/or theoretical knowledge | 83.3 | ✓ |
| Research of direct or potential impact, across one or more lines of investigation, demonstrates a logical trajectory (i.e. projects successively build on completed research) | 8.3 | 🗴 |
| Research plan is innovative (e.g. generates novel methods, models, data or other knowledge that addresses a noted gap | 87.5 | ✓ |
| Research program includes higher and lower risk lines of investigation to balance feasibility | 37.5 | 🗴 |
| Program of research aligns with high-level organizational research priorities | 8.3 | 🗴 |
| Program of research considers sex, gender and intersectional factors as relevant in research aims, rationale, participants, analyses, and also among research team, staff and trainees | 29.2 | 🗴 |
| Challenges to Research Productivity |  |  |
| Qualitative description by researcher of challenges faced and mitigating strategies applied (e.g. no funding for administrative or research assistant so used an undergraduate volunteer to help prepare research funding applications) | 50.0 | ✓ |
| To supplement qualitative description of challenges, objective description of infrastructure provided by organization so that research institute understands potential impact on progress | 33.3 | 🗴 |
| Team/Open Science |  |  |
| Where it would benefit the research topic or program, evidence of local, regional, national or international team science (e.g. formation of clinical network or inter-/multi-disciplinary research group) | 54.2 | ✓ |
| Evidence of collaboration or multidisciplinary research through participation as a co-investigator or mentor on other research teams | 75.0 | ✓ |
| For research directly related to patient care, co-production of research with those outside academia (e.g. policy-makers, patients/family, public) | 29.2 | 🗴 |
| If relevant registries are available, registration of planned research (e.g. trials, literature syntheses) | 20.8 | 🗴 |
| Open publication, and where relevant to type of research, open sharing of research outputs (e.g. data sets, software, code, biological materials, tools) | 37.5 | 🗴 |
| Funding |  |  |
| Evidence of research independence (e.g. PI or co-PI) OR of a fundamental role on a research team (e.g. biostatistician, qualitative researcher) with peer-reviewed research funding | 87.5 | ✓ |
| A balance of peer-reviewed funding from Tri-Council or other similar sources with funding from other sources | 20.8 | 🗴 |
| Attempts to capture peer-reviewed research funding for proposals with merit (evidence of positive feedback, ranking or scores) | 37.5 | 🗴 |
| Attempts to capture non-peer-reviewed research funding to supplement or fill gaps in peer-reviewed research funding (e.g. number of submitted applications) | 16.7 | 🗴 |
| Attempts to capture peer-reviewed, competitive salary support such as New Investigator or Chairs (e.g. number of applications for nominations) | 8.3 | 🗴 |

| Innovations |  |  |
| --- | --- | --- |
| Evidence of research outputs/products relevant to type of research (researcher can choose from):   - Commercialization of technology (e.g. software, drugs, devices), launch of companies, invention disclosures, patent applications, issued patents, licenses, etc - Creation of cohorts or registries - Clinical tests, algorithms or statistical models - Validated questionnaires or instruments - Contribution to policies, standards, guidelines or programs - Novel theory, model or framework - Novel research approaches or methods - Other forms of creative achievement or outputs relevant to discipline (e.g. physical simulation, performance art) | 62.5 | ✓ |
| Dissemination – Publications |  |  |
| Quality of the content of publications as judged by a peer reviewer or panel, in part based on rationale provided by researcher of importance to field | 87.5 | ✓ |
| Number of peer-reviewed journal publications of high quality (as judged by a peer reviewer or panel, in part based on rationale provided by researcher of importance to field) | 45.8 | 🗴 |
| Apart from peer-reviewed journal publication, evidence of other types of reports, relevant to type of research, to disseminate research findings (e.g. books, book chapters, preprints, editorials, commentaries, guidelines, task force reports) | 29.2 | 🗴 |
| Dissemination – Presentations, Social Media, Creative Approaches |  |  |
| Recognition at academic or professional society meetings (e.g. invited presentations or workshops, award for poster or presentation) | 54.2 | ✓ |
| Evidence of dissemination activities relevant to type of research (e.g. presentations or meetings with healthcare professionals, policy-makers, community groups or general public; social media, interviews) | 29.2 | 🗴 |
| Evidence of Impact |  |  |
| Evidence of impact relevant to type of research and career stage. Researcher can choose from the following or specify other impact:   - Researcher reputation (e.g. leadership role in local, provincial, national or international committees, organizations, or conference steering groups) - Distinctions, credentials, honours and awards, funded and unfunded - Leadership or co-leadership of clinical translation through Phase 1 to 3 clinical trials - Contributions to organizational, government or health system policy development or changes - Demonstrated use of research outputs by other researchers, clinicians, organizations, government, patients/families - Social, cultural, population, environmental or economic returns from use of research outputs/products and associated outcomes | 45.8 | 🗴 |
