## Supplemental Appendix 8 for "DORA-compliant measures to assess research quality and impact in biomedical institutions: review of published research, international best practice and Delphi survey"

**APPENDIX 8. Perceived pros and cons of measures that did not achieve consensus**

| Measures  [rated important by % panelists] | Exemplar comments |
| --- | --- |
| Relevance of Research Program | |
| Research of direct or potential impact, across one or more lines of investigation, demonstrates a logical trajectory (i.e. projects successively build on completed research) [8.3] | PRO  None offered  CON   - Not relevant to early career researchers who have not yet generated a body of research - Might compel researchers to avoid branching out into to explore new ideas if they follow a rigid path |
| Research program includes higher and lower risk lines of investigation to balance feasibility [37.5] | PRO  Makes for a productive research program  CON  Balance of higher- to lower-risk research should be left to the individual researcher |
| Program of research aligns with high-level organizational research priorities [8.3] | PRO  None offered  CON  Organizational priorities subject to change with trends and leadership. May stifle creativity and innovation |
| Program of research considers sex, gender and intersectional factors as relevant in research aims, rationale, participants, analyses, and also among research team, staff and trainees [29.2] | PRO  Essential components of all research programs  CON  None offered |
| Challenges to Research Productivity | |
| To supplement qualitative description of challenges, objective description of infrastructure provided by organization or sponsors so that research institute understands potential impact on progress [33.3] | PRO  Will unveil problems with Institutional infrastructure  CON  Qualitative should suffice |
| Team/Open Science | |
| For research directly related to patient care, co-production of research with those outside academia (e.g. policy-makers, patients/family, public) [29.2] | PRO  Ability to secure and lead a team is evidence that they can move their goal forward in an effective manner  CON  A "one size fits all" approach has unintended consequences |
| If relevant registries are available, registration of planned research (e.g. trials, literature syntheses) [20.8] | PRO  Becoming more important  CON  None offered |
| Open publication, and where relevant to type of research, open sharing of research outputs (e.g. data sets, software, code, biological materials, tools) [37.5] | PRO  Sharing of data is an important component of all research  CON  Grants in my area don't cover the costs of this |
| Funding | |
| A balance of peer-reviewed funding for Tri-Council or other similar sources with funding from other sources [20.8] | PRO  Evidence of peer-review at the 'highest' level of funding within Canada  CON  Not sure it matters where the money comes from |
| Attempts to capture peer-reviewed research funding for proposals with merit (evidence of positive feedback, ranking or scores) [37.5] | PRO  If quality applications are not being funded this should be recorded. Very important metric especially related to EDI.  CON  Continual close-but-not-quite-funded does not run a lab |
| Attempts to capture non-peer-reviewed research funding to supplement or fill gaps in peer-reviewed research funding (e.g. number of submitted applications) [16.7] | PRO  None offered  CON  Will encourage quick, not well thought out grant applications |
| Attempts to capture peer-reviewed, competitive salary support such as New Investigator or Chairs (e.g. number of applications for nominations) [8.3] | PRO  None offered  CON  The institution nominates for most of these. If that isn't done equitably the whole question is meritless |
| Dissemination – Publications | |
| Number of peer-reviewed journal publications of high quality (as judged by a peer reviewer or panel, in part based on rationale provided by researcher of importance to field) [45.8] | PRO  Publications remain one of the most impactful metric of a PI's success  CON   - Number does not mean anything unless scaled to the size of the group - Is there any distinction for authorship position? |
| Apart from peer-reviewed journal publication, evidence of other types of reports, relevant to type of research, to disseminate research findings (e.g. books, book chapters, preprints, editorials, commentaries, guidelines, task force reports) [29.2] | PRO  Broadens out the concept of publications and may apply to many disciplines  CON  None offered |
| Dissemination – Presentations, Social Media, Creative Approaches | |
| Evidence of dissemination activities relevant to type of research (e.g. presentations or meetings with healthcare professionals, policy-makers, community groups or general public; social media, interviews) [29.2] | PRO  Needs to be judged in the context of the PI's research objectives  CON  None offered |
| Evidence of Impact | |
| Evidence of impact relevant to type of research and career stage. Researcher can choose from the following or specify other impact:   - Researcher reputation (e.g. leadership role in local, provincial, national or international committees, organizations, or conference steering groups) - Distinctions, credentials, honours and awards, funded and unfunded - Leadership or co-leadership of clinical translation through Phase 1 to 3 clinical trials - Contributions to organizational, government or health system policy development or changes - Demonstrated use of research outputs by other researchers, clinicians, organizations, government, patients/families - Social, cultural, population, environmental or economic returns from use of research outputs/products and associated outcomes [45.8] | PRO  Important from the stand-point of not only impact, but also career development, evidence of leadership potential, being a team-player, giving-back, etc.  CON   - Seems to leave out the basic science people - Judgment of this will be pretty subjective |
